# What Embedding-Based Guideline–Literature Drift Measures (and What It Doesn’t): A Proof-of-Concept and Mechanistic Dissection of TIDE in Orthopedic Surgery

**DOI:** 10.64898/2026.09.28.26364191

**Authors:** Colby Grames, Paul Zakarian, Christopher Franquemont, Andrew Cabrera, Joseph Elsissy

## Abstract

**Purpose:** We introduce the Temporal Index of Divergent Evidence (TIDE), a metric that evaluates how the topic center of gravity of a corresponding corpus of literature aligns with, or shifts relative to, a recommendation’s semantics over time.

**Methods:** TIDE computes annual mean cosine similarity using MedCPT embedding vectors between a recommendation and corresponding PubMed records binned by year (2000-2024), testing for monotonic temporal trend (Kendall τ). Three recommendations with expected divergent, stable, and convergent trajectories were examined: two-stage exchange for periprosthetic joint infection (PJI), emergent fasciotomy for acute compartment syndrome (ACS), and non-operative management of distal radius fractures (DRF) in adults ≥ 65 years. The signal was characterized by an additive per-token attribution, an isolated vs. in-context analysis, and an adversarial minimal-pair test.

**Results:** PJI diverged (τ = −0.84, year-shuffle p = 0.001), DRF converged (τ = +0.42, p = 0.005), and ACS showed no detectable trend (τ = −0.07, p = 0.66). The two significant directions (PJI, DRF) held under matched input [CLS] pooling, while ACS remained near zero. Publication volume baselines did not distinguish the three trajectories (publication count was universally positive, τ = +0.77 to +0.97). A reversed-meaning minimal-pair control showed little cosine sensitivity to reversal. Paper-level TF-IDF recovered the qualitative three-case pattern.

**Conclusion:** Embedding-based drift analysis is a proof-of-concept candidate pre-triage signal that tracks topical alignment, not evidentiary direction; superiority over TF-IDF remains unproven. The present study is hypothesis-generating, showing potential use of embedding representations as an analytical unit rather than simply for retrieval.

## 1 INTRODUCTION

The expansion of biomedical literature in recent years has made it increasingly difficult to assess current clinical practice guidelines. While guidelines can be critical and necessary tools for standardizing care, at best they serve only as static summaries of an ever-growing body of evidence. Shekelle et al. found that approximately half of clinical guidelines become outdated within 5.8 years of publication, with only 90% still valid at 3.6 years. [1] The same decay pattern holds at every level: approximately one in five individual recommendations are out of date within three years, while nearly a quarter of systematic reviews require updating within two years (median survival of 5.5 years). [2,3] The growing volume of clinical research further increases the burden of keeping recommendations current. [4] In orthopedic surgery, where the American Academy of Orthopedic Surgeons is responsible for maintaining a plethora of guidelines and recommendations across different subspecialties, the supply of human expertise is being vastly exceeded by demand.

The tension lies not only in guidelines becoming outdated, but also in identifying which guidelines have become outdated and need prompt updating-the problem of triage. Clinicians perceive this friction most prominently; physician surveys have long documented meaningful disagreement with published guidelines, and the rapid deterioration of guideline validity can compound uncertainty at the point of care. [1,5] Identifying outdated guidelines often relies on periodic literature searches and expert review. [6] Structured prioritization methods have been described but remain effort-intensive and periodic, so meaningful shifts in evidence go unrecognized for prolonged periods.

The management of periprosthetic joint infection demonstrates this problem in real time.

Two-stage exchange is commonly used for PJI, while single-stage exchange and, in selected early or acute infections, debridement with implant retention are alternatives. [7–11] Efforts have been undertaken to address the problem by employing natural language processing and text-mining models to find and condense the relevant literature as it is published. [12] However, these models primarily rely on predefined search strategies and supervised classification of study records against human labels, rather than measuring how the literature’s topical alignment to a specific recommendation changes over time. [13] Vector embedding-derived methods have begun to enter the medical domain, retrieving literature relevant to guideline passages or ranking trials for review updates. [14,15] Unsupervised embeddings have previously been used to characterize how the thematic structure of a literature shifts over time. These studies analyze thematic change rather than alignment to a fixed recommendation. [16,17] Semantic-distance methods have supported guideline-update retrieval, and embedding-based guideline-document similarity has been examined over time in regulatory assessment reports. [18,19] Here, we examine whether the literature’s topical center of gravity shifts relative to a recommendation’s wording over time. This work extends these methods to an anchor-relative, unsupervised, longitudinal setting.

To address this concern, we have developed the Temporal Index of Divergent Evidence (TIDE), a pilot framework that measures semantic drift between a fixed recommendation and its year-binned literature using biomedical text embeddings. [20] Rather than linking citations and counting studies, TIDE evaluates how the topical center of gravity of the corresponding literature aligns with, or shifts relative to, a recommendation’s wording over time. We position TIDE as a candidate pre-triage signal that could help allocate valuable expert time to recommendations that may warrant earlier human review. Whether topical drift can, at scale, identify which recommendations may warrant examination remains open and unresolved. Generative methods of analysis have shown tendencies to fabricate and miscite guideline evidence; thus, the pipeline is deliberately deterministic and does not use generative language models in its analysis. [21]

The purpose of the present study is to assess whether embedding-based semantic tracking can recover the expected directions of topical drift (divergence, convergence, and stability); whether the observed signals are robust to procedural variation; and to perform an exploratory analysis of the interpretability of the present method.

## 2 METHODS

### 2.1 Study Design

TIDE (Temporal Index of Divergent Evidence) measures the annual mean cosine similarity between an orthopedic recommendation anchor and the contemporary published literature in a learned (MedCPT) embedding space. It then assesses whether the calculated similarity exhibits a monotonic temporal trend. We retrospectively applied TIDE to three recommendations with expected trajectories during the current century (Figure 1): treatment of periprosthetic joint infection with two-stage exchange arthroplasty (expected divergence with the growing interest in single-stage and Debridement, Antibiotics, and Implant Retention (DAIR) techniques), treatment of acute compartment syndrome (ACS) with emergent fasciotomy (expected stability, a long-settled known practice), and operative versus non-operative management of distal radius fractures (DRF) in patients 65 or older (expected convergence, evidence increasingly supports nonoperative management for long term patient reported outcomes).

**Fig 1.**
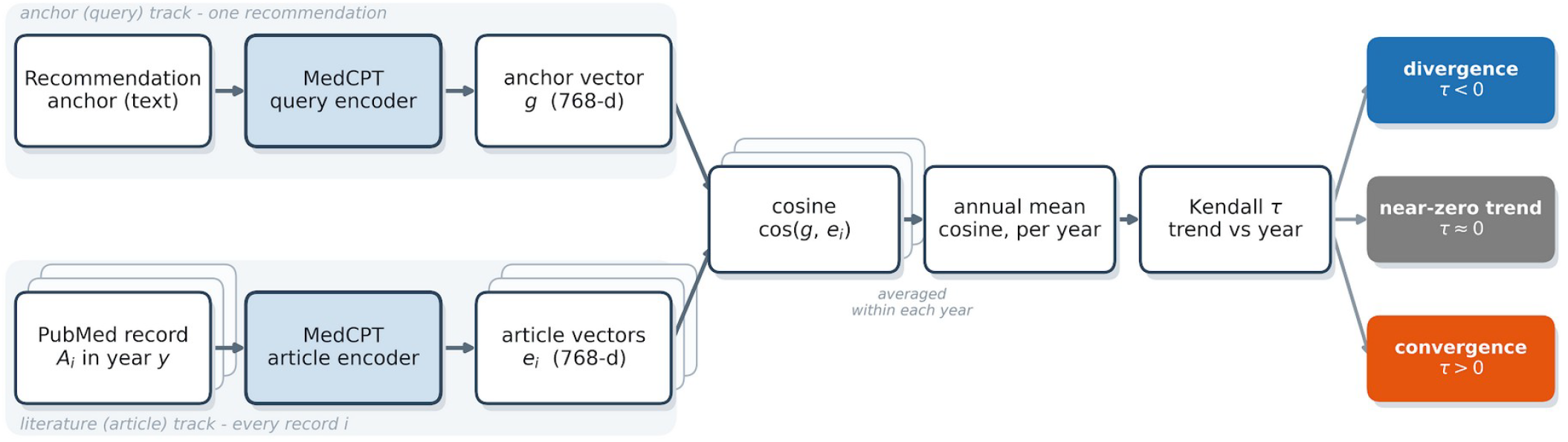
The TIDE pipeline. Each recommendation anchor is embedded with the MedCPT query encoder and each year’s PubMed records with the article encoder; the annual mean cosine similarity is tested for a monotone temporal trend (Kendall τ), classifying the relationship as diverging (τ < 0), little monotonic association (τ ≈ 0), or converging (τ > 0).

### 2.2 Recommendation Anchors

Each recommendation was compressed to its clinical essence, removing preambles, complex grammatical structures, and other non-essential phrasing. All anchors were subjected to the same compression criteria and refinement by the principal investigator.

PJI and ACS anchors are representative statements of established clinical practice rather than verbatim AAOS recommendations. The DRF anchor was adapted from a corresponding 2020 AAOS recommendation, with modifications only made to clarify the underlying assertion. [22] Table 1 lists the exact anchor phrases used in the analysis.

**Table 1.** Recommendation anchors and literature corpora. The verbatim text embedded as each recommendation anchor, its source, the abbreviated PubMed query defining the annual corpus, the number of records (n), and the years analyzed.

| Case | Anchor text embedded (verbatim) | Source | PubMed query (abbreviated) | n | Years |
| --- | --- | --- | --- | --- | --- |
| PJI | “Chronic periprosthetic joint infection should be treated with two-stage exchange arthroplasty.” | Representative statement (no single verbatim AAOS recommendation) | (periprosthetic OR prosthetic joint infection OR PJI) AND (two-stage OR 2-stage OR exchange arthroplasty OR revision) | 2,405 | 2008–2024 |
| ACS | “Acute compartment syndrome should be treated with emergent fasciotomy.” | Representative statement | (acute compartment syndrome OR compartment syndrome) AND (fasciotomy OR decompression OR surgical) | 1,463 | 2000–2024 |
| DRF | “Operative treatment for geriatric patients aged 65 years and older does not lead to improved long-term patient-reported outcomes compared to non-operative treatment for distal radius fractures.” | Adapted AAOS 2020 CPG recommendation | (distal radius fracture OR wrist fracture) AND (elderly OR geriatric OR older OR aged OR osteoporotic) | 636 | 2000–2024, except 2001 |

*All records in English, 2000–2024, with $\geq 5$ publications/year retained. Anchors embedded with the MedCPT query encoder; records with the MedCPT article encoder. PJI/ACS anchors are representative statements; DRF is adapted from the AAOS 2020 recommendation. Exact queries are in anchors.json.*

### 2.3 Literature Corpus

For each anchor, we constructed a PubMed query (Table 1) to maximize coverage of relevant records while minimizing off-topic Records. Records were limited to English-language records published between 2000 and 2024. Calendar years for a given anchor were excluded from analysis if fewer than five records were found. The final analytical corpus consisted of 2,405 PJI, 1,463 ACS, and 636 DRF records, respectively. Titles were concatenated to their respective abstracts; title-only records were retained. Reviews, systematic reviews, meta-analyses, case reports, editorials, letters, and other non-primary research publication types were excluded as specified in anchors.json.

### 2.4 Embeddings and Similarity

MedCPT is a set of two encoders (query and article) trained on pairs of PubMed queries and retrieved articles. We used the query encoder for the recommendation statement, and embedded the records with the article encoder. This computation yielded a 768-dimensional vector per token contextualized by internal attention. Attention-mask-selected per-token vectors, including special tokens, were then averaged into a single 768-dimensional vector for the recommendation and for each record; finally, the resulting mean vectors were then L2-normalized for standardization. For each record, the cosine similarity to the corresponding recommendation anchor was computed, then pooled into year-binned mean cosine similarity scores.

### 2.5 Trend Statistics

The primary metric was the Kendall rank correlation τ between calendar year and the annual mean cosine, with the Theil-Sen slope reported as the corresponding effect size. A negative τ denotes “divergence” (records in the surrounding literature decreasing in cosine similarity in MedCPT’s learned embedding space), a positive τ denotes “convergence”, and a τ near zero denotes little monotonic association, not necessarily evidence of stability.

### 2.6 Inference and Null Models

Uncertainty was quantified with nonparametric intra-year bootstrapping (resampling records within each included year; 1,500 resamples for ranges; 1,000 resamples for sign preservation). We report the central 95% conditional resampling range on τ and the percentage of replicates that preserved its sign. These are not calibrated confidence intervals and can exclude the observed τ. Record-year labels were shuffled before reapplying the annual-count floor (1,000 iterations), where the probability of a |τ| at least as large as the observed value was recorded as a null test. A phase-randomized null used 5,000 replicates and treated retained annual bins as equally spaced, including the DRF gap. All stochastic checks used seed 0. Additional robustness checks fit a full-data ordinary least squares regression of record-level cosine on year, with standard errors clustered by year, and repeated the trend under leave-one-year-out resampling.

### 2.7 Pooling Robustness

MedCPT natively represents each body of text with a special classification ([CLS]) token vector rather than by mean over all tokens. We thus recomputed each annual-mean-cosine trend under [CLS] pooling on the same records, anchors, years, and single-string 512-token inputs, changing only the pooling operation.

### 2.8 Baselines

To examine whether the indexed similarity trends reflected publication growth rather than semantic shift, we computed the annual publication count and volume of higher-evidence studies (comparative studies, clinical trials, randomized controlled trials, or multicenter studies, as indicated by declared PubMed type). This potential confounder was further probed by computing partial Kendall rank correlation between year and binned annual mean cosine similarity while controlling for annual record volume, and fit an ordinary least squares model of annual mean cosine similarity on a year and log-transformed volume relationship. Because annual volume and calendar year were very strongly collinear in the present corpora (τ = 0.77 − 0.97), joint covariate estimation is not fully resolvable. The direct comparison and adjustment for drift versus volume trends therefore remained the primary confound test, with the partial correlation reported as a supporting check. Post hoc matched TF-IDF comparisons varied annual document versus paper-level IDF fitting and scoring; paper-level cosines were averaged within year. Fitting included the anchor, English stopword removal, minimum document frequency 2, and sublinear term frequency. Evidence-weighted analyses used the maximum matching publication-type weight, with a minimum of 1; weights are specified in data/sensitivity/methods.json.

### 2.9 Interpretability

In order to understand individual key terms’ impact on the observed phenomenon, we decomposed each record’s cosine similarity into per-term contributions and tracked how each term’s contribution changed over time. The cosine of a mean-pooled embedding set is an exact sum of per-token contributions given by: cos(g, e) = Σ_i_ (g · h_i_) / (N‖e‖), where g is the unit-normalized anchor vector, N is the number of tokens in the record, h_i_ are the contextual token vectors, and e is their unnormalized mean. We further examined each term’s isolated embedding (the term embedded alone) alongside its contextualized embedding and constructed minimal pairs that use identical sets of words rearranged to reverse the contextual meaning, in order to test whether the embedding encodes evidentiary direction. The study will refer to these checks as the exact cosine attribution and minimal pair probes, respectively. The exact decomposition requires mean pooling; the minimal-pair probe is model-agnostic. Token-level analyses used 400 token inputs; sampling settings are specified in README.md. Finally, as an exploratory geometric analysis, principal component analysis was applied to the per-year record centroids of the PJI corpus; the resulting axes were labeled by mapping the clinical terms that most closely associated with each axis. A post hoc specificity check scored each of the three corpora against all three anchors.

### 2.10 Statistical Software and Reproducibility

Analysis was performed in Python (3.13; NumPy, SciPy, scikit-learn) using the Hugging Face Transformers implementation of MedCPT. [20] Tests were two-sided with nominal significance at p < 0.05, without multiplicity adjustment. Frozen queries, PMID mappings, vectors, numerical outputs, and analysis scripts are versioned at https://github.com/Colbstang/TIDE. Text-dependent reproduction requires separately retained frozen normalized records; the original search timestamp and raw XML were not archived.

Generative AI assistance: OpenAI ChatGPT/Codex assisted literature searching, methodological review, code development, reproducibility checks, and manuscript copyediting and revision. These tools were not components of TIDE scoring. The authors retain responsibility for the analyses, references, interpretation, and final manuscript.

## 3 Results

Thee present study analyzed 4,504 case-record observations (4,503 distinct PubMed IDs; 29 title-only records) across three recommendation anchors: two-stage periprosthetic joint infection treatment (PJI; n = 2,405, 17 years, 2008-2024), emergent fasciotomy for acute compartment syndrome (ACS; n = 1,463; 25 years; 2000-2024), and non-operative management of distal radius fractures in patients ≥ 65 years (DRF; n = 636; 24 years; 2000-2024, excluding 2001). Only binned years with ≥ 5 records were analyzed; therefore, PJI was restricted to 2008-2024 by this annual-count threshold. Corpus density varied by anchor; PJI had a median record count of 94 [range 6-364], ACS 61 [17-109], and DRF 19 [5-67]. Corpus volume also grew markedly over the studied range; PJI annual volume, for example, rose from 9 records/yr in 2008 to 364 records/yr in 2024.

The three anchor statements, created to capture perceived change in literature and practice, showed a mean annual cosine similarity that moved in the expected direction in every case (Table 2, Figure 2). Measured cosine similarity to the PJI anchor steadily declined between 2008 and 2024 (τ = −0.84; Theil-Sen slope = −9.7×10^−4^/yr; year shuffle p = 0.001; phase-randomized p = 0.0002; sign preserved in 100% of intra year bootstrap iterations), the DRF anchor converged (τ = +0.42; Theil-Sen slope = 3.6×10^−4^/yr; phase-randomized p = 0.012; year shuffle p = 0.005; sign preserved in 98% of intra year bootstrap iterations), and ACS showed no detectable monotonic trend (τ = −0.07; Theil-Sen slope = −0.6×10^−4^/yr; year shuffle p = 0.66; sign preserved in 71% of intra year bootstrap iterations). The two directional cases (PJI, DRF) held under matched-input [CLS] pooling (PJI τ = −0.57; DRF τ = +0.40), while ACS remained near zero (τ = +0.03). The pattern also held under full-data robust clustering OLS (PJI p < 0.0001; DRF p = 0.0032; ACS p = 0.75), and leave-one-year-out resampling (Table 2).

**Table 2.** Semantic-drift signal and robustness battery for the three index recommendations (Kendall τ on annual mean cosine). Bold τ values have nominal year-shuffle p < 0.05; ranges are conditional resampling intervals.

| Case | Kendall $\tau$ | 95% conditional range | Slope ($\times 10^{-4}/\text{yr}$) | Year-shuffle $p$ | Boot same-sign | Matched CLS $\tau$ | Full-data OLS | Leave-one-yr-out $\tau$ | Evidence-wt $\tau$ | Expected → Observed |
| --- | --- | --- | --- | --- | --- | --- | --- | --- | --- | --- |
| PJI | **-0.838** | [-0.82, -0.53] | -9.7 | **0.001** | 100% | -0.574 | -11e-4/yr, $p < 0.0001$ | [-0.87, -0.82] | -0.779 | divergence → divergence ✓ |
| ACS | -0.067 | [-0.31, +0.17] | -0.6 | 0.66 n.s. | 71% | +0.033 | -0.39e-4/yr, $p = 0.746$ | [-0.12, +0.01] | -0.060 | expected stability → no detectable trend |
| DRF | **+0.420** | [+0.04, +0.47] | +3.6 | **0.005** | 98% | +0.399 | +3.99e-4/yr, $p = 0.0032$ | [+0.375, +0.478] | +0.428 | convergence → convergence ✓ |

CLS = matched-input [CLS]-pooling recompute; OLS = full-data cluster-robust regression; LOO = leave-one-year-out. Conditional resampling ranges may exclude the observed $\tau$ . All values use the primary anchors. Query sensitivity (31 Aug 2026; 5 variants/case): PJI 100% negative, DRF 100% positive, with nominal $p < 0.05$ in 4/5 and 3/5, respectively; ACS had no significant trend under five labels representing four unique queries.

**Fig 2.**
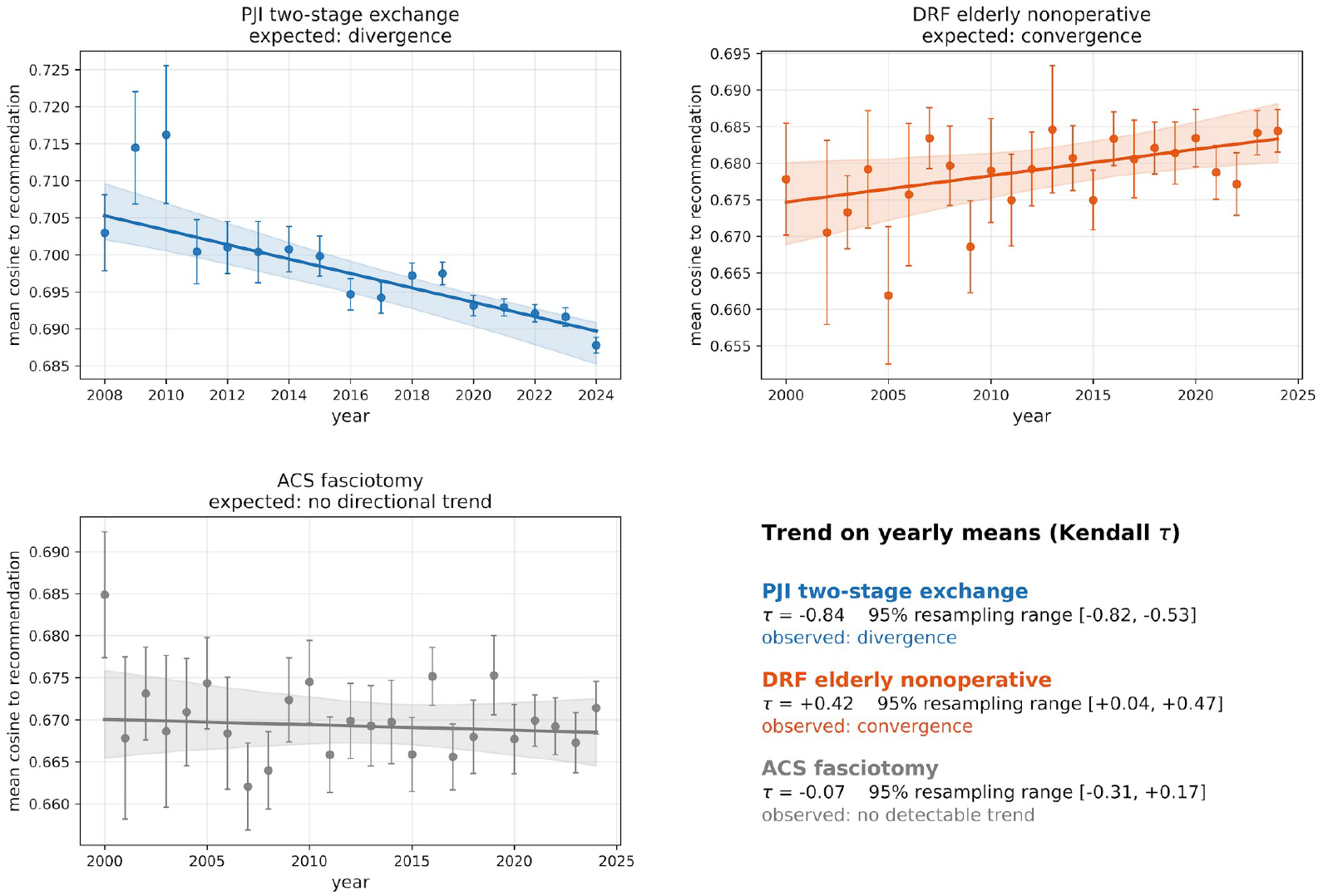
Semantic-drift trajectories for the three index recommendations. Annual mean cosine to the recommendation anchor (points, ±1 SE) with Theil–Sen trend and conditional within-year bootstrap 95% pointwise fitted-line band (shaded). PJI two-stage exchange diverges and elderly DRF nonoperative management converges (top); ACS fasciotomy is a flat working negative control (bottom).

To ascertain whether the present trends were artifacts of the method rather than real, interpretable signals, two avenues were examined. First were null controls and query-sensitivity checks: year-shuffle permutation gave nominal p ≤ 0.005 for the PJI and DRF anchors but not for the ACS anchor (p = 0.66); a separately dated query sensitivity reanalysis (31 Aug 2026) found negative PJI and positive DRF trends in all fivve variants, with nominal p < 0.05 in 4/5 and 3/5, respectively. ACS had no nominally significant trend under fivve labels representing four unique query definitions. Second, the confound baselines (Table 3.): annual publication count and annual higher evidence study count were uniformly positive over all three corpora (publication count τ: +0.967 PJI, +0.768 ACS, +0.854 DRF; evidence volume τ: +0.711 PJI, +0.289 ACS, +0.544 DRF) and thus cannot distinguish a divergent case from a convergent. Consistent with this result, the correlation between annual mean cosine and annual volume was itself divergent in sign across cases (τ = −0.83 PJI, −0.07 ACS, +0.40 DRF), suggesting against but not ruling out publication-volume confounding. Thee expected directions were preserved under partial correlation controlling for annual volume (partial τ = −0.23 PJI, −0.02 ACS, +0.16 DRF), though atteenuated; year coefficcients in full OLS adjustment could not separate volume and calendar year due to high collinearity.

**Table 3.** Publication-count and evidence-volume baselines.

| Case | TIDE $\tau$ | Publication-count $\tau$ | Evidence-volume $\tau$ |
| --- | --- | --- | --- |
| PJI (divergence) | **-0.838** | +0.967 | +0.711 |
| ACS (flat) | -0.067 | +0.768 | +0.289 |
| DRF (convergence) | **+0.420** | +0.854 | +0.544 |

*Publication-count and evidence-volume $\tau$ are uniformly positive and cannot separate divergence, convergence, and flat trajectories. Bold = significant TIDE trend*

A Lexical (TF-IDF) baseline was computed as a comparator. The annual document baseline on matched inputs reproduced the PJI and DRF directions (τ = −0.94, +0.14) but flagged the ACS case as strongly diverging (τ = −0.72) and inverted the DRF direction under the historical shorter DRF anchor (which changes the clinical assertion) (τ = −0.57). However, the paper-level IDF fitting with annual means of paper-level cosines recovered the qualitative three-case pattern (PJI τ = −0.44; ACS τ = −0.02; DRF τ = +0.63), embedding superiority in this regard was not established.

The cosine of a mean-pooled embedding vector can be decomposed into contributions from individual tokens; we thus tracked which terms contributed to divergence in the PJI case (Figure 3). Contribution from the two-stage related vocabulary (two-stage, reimplantation, debridement) peaked around 2010-2014 and then diminished, while the contribution from other terms related to single-stage or DAIR procedures (one-stage, single-stage, DAIR) rose from near 0 at the beginning of the window but remained below two-stage at the end of the window. As an exploratory geometric analysis, principal component analysis (PCA) of per-year PJI record mean pooled centroids (Figure 4) showed the corpus moving along a mapped interpretable axis (PC1, 48% of variance) running from diagnosis/microbiology to treatment-procedure content, with the 2008 to 2024 trajectory showing a steady trend along the PC1 (Kendall τ of year vs PC1 score, τ = +0.94) axis. The PCA analysis is presently exploratory; axes are data-derived, and no significance is claimed.

**Fig 3.**
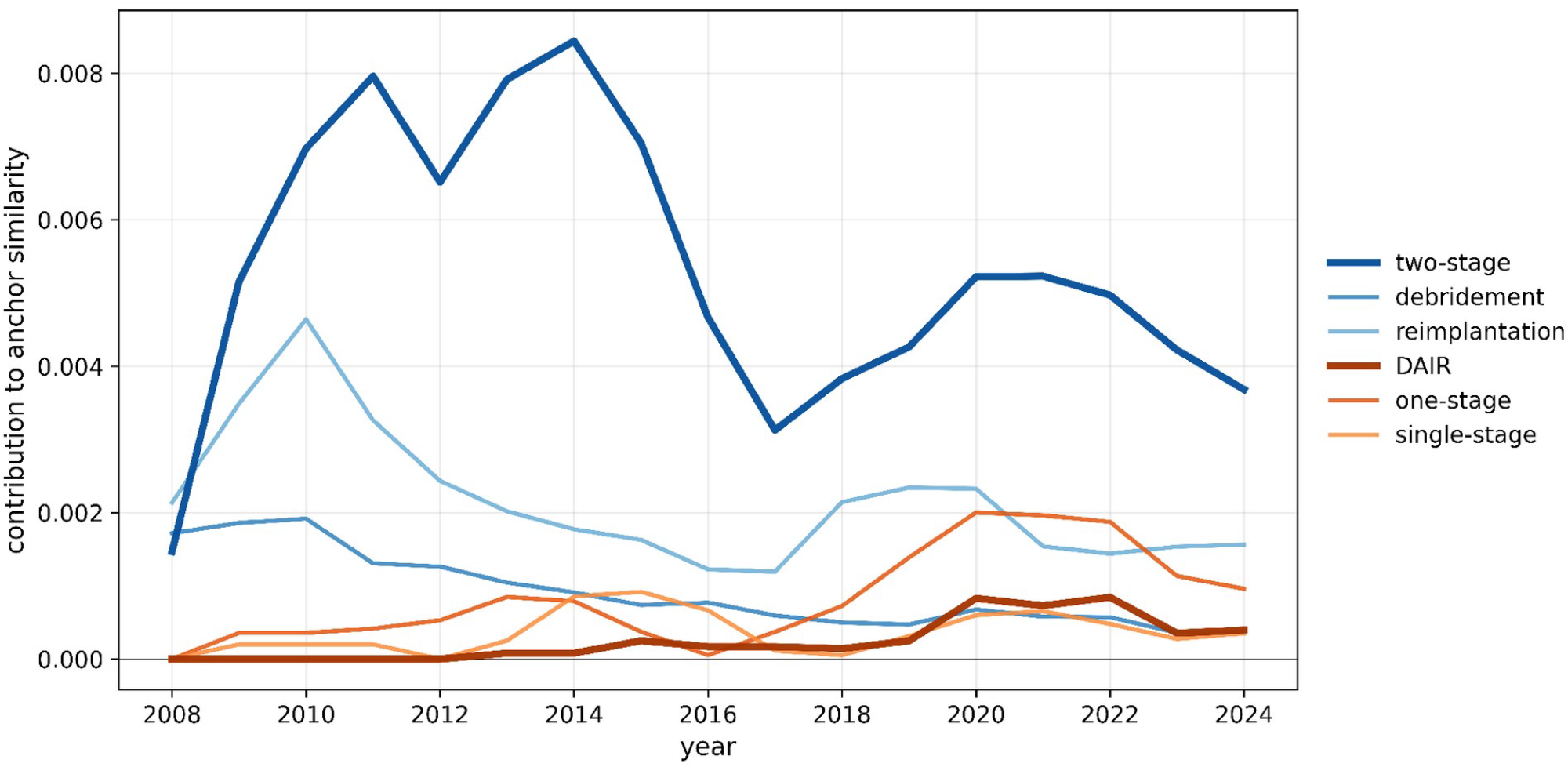
What the PJI signal is made of. Per-term contribution to the guideline–abstract cosine over time, from the exact per-token decomposition: two-stage exchange vocabulary recedes while single-stage/DAIR terms rise, descriptive contributors to the PJI divergence. Up to 40 PJI records/year were sampled (seed 0), with 400-token inputs and centered three-bin smoothing; the six terms do not exhaust the cosine score.

**Fig 4.**
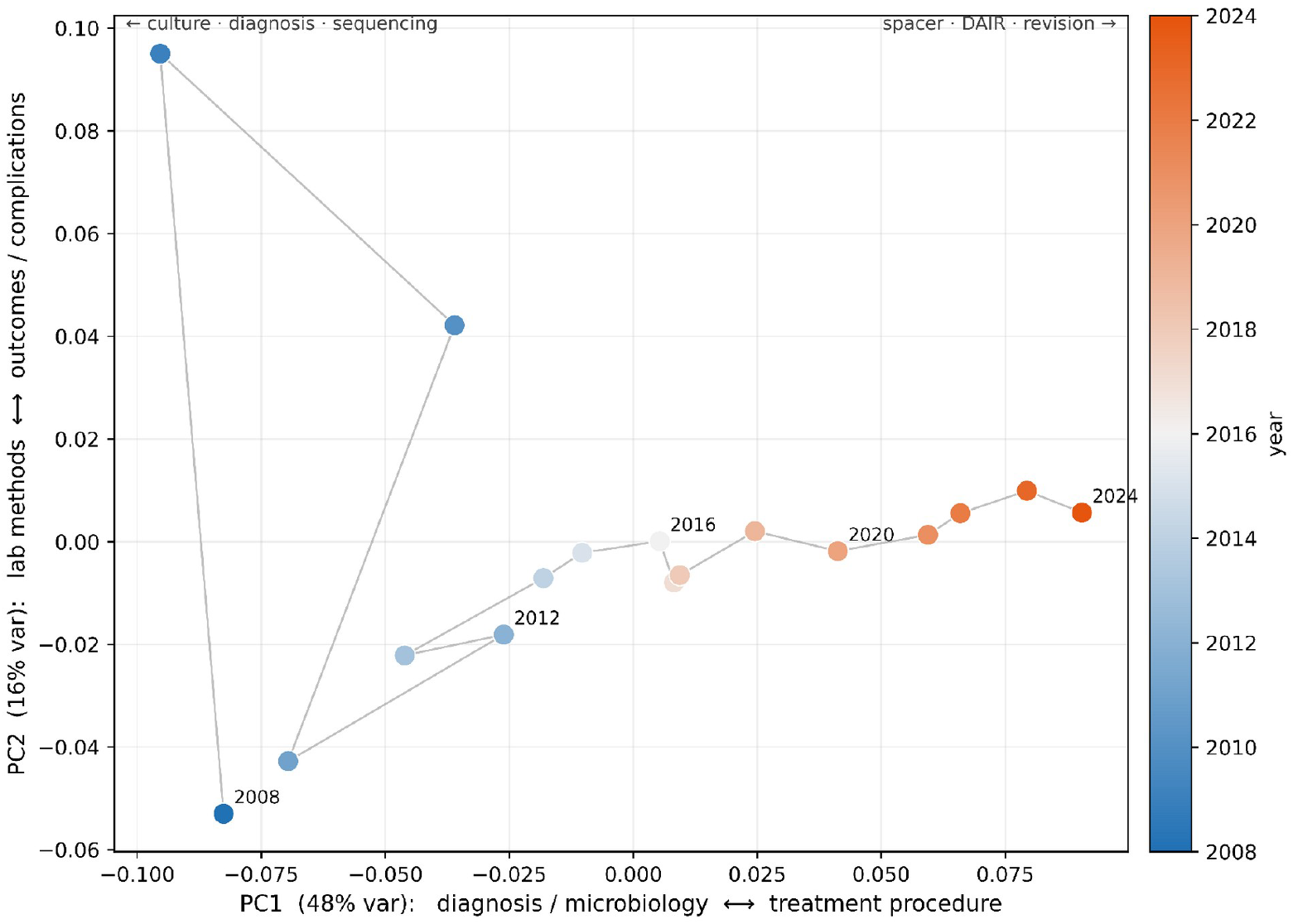
The PJI literature drifts along an interpretable axis of MedCPT space (exploratory). PCA of per-year abstract centroids; PC1 (48% of variance), named by post hoc clinical-term projections, runs from diagnosis/microbiology to treatment-procedure, and the year trajectory (2008→2024) moves along it. Axes are data-derived; no significance is claimed.

The signal measured was then tested to determine whether embeddings can read the evidentiary direction or only topical similarity (Table 4). For PJI and DRF, two sentences used an identical set of words with the claimed direction reversed (ex., “Nonoperative management was superior to operative fixation” vs. “operative fixation was superior to nonoperative management”). Reversing the claim changed the MedCPT cosine to the anchor statement by less than 0.003 (PJI: 0.760 vs 0.757; DRF: 0.799 vs 0.798) and changed TF-IDF cosine by 0.000 in both cases. These two probes did not establish reliable discrimination of reversed meaning.

**Table 4.** Minimal-pair probe: topic versus evidentiary direction. Cosine to the recommendation anchor for paired statements with identical wording but reversed meaning. The DRF probe uses the shorter anchor in data/sensitivity/methods.json, not the primary anchor.

| Statement (same bag of words, meaning flipped) | TIDE cos | TF-IDF cos |
| --- | --- | --- |
| DRF — “Nonoperative management was superior to operative fixation...” | 0.799 | 0.185 |
| DRF — “Operative fixation was superior to nonoperative management...” | 0.798 | 0.185 |
| PJI — “Two-stage exchange was superior to single-stage exchange...” | 0.760 | 0.502 |
| PJI — “Single-stage exchange was superior to two-stage exchange...” | 0.757 | 0.502 |

*Flipping the meaning changes the score by $< 0.003$ (TIDE) and $0.000$ (TF-IDF); neither method reliably distinguished stance in these probes — the signal tracks topic, not stance.*

## 4 DISCUSSION

In the present study, we investigated whether analysis of embedding representations of records related to a given anchor statement could reconstruct expected temporal trends in medical literature. We recovered the expected pattern in three selected cases: periprosthetic joint infection (PJI), distal radius fracture (DRF), and acute compartment syndrome (ACS). The directional cases (PJI and DRF) were robust to year shuffle permutation, conditional intra-year resampling, and matched input CLS pooling. Furthermore, we found that publication volume trends differed from TIDE trends, but adjusted estimates were attenuated and imprecise, and did not rule out confounding factors entirely. Paper-level TF-IDF also recovered the qualitative three-case pattern. The authors interpret these results as evidence that the method indexes semantic drift in the literature over time.

Confidence that this signal reflects a real shift, rather than an artifact of a single analytical choice, rests on the convergence of three complementary analyses. The trend statistic first locates the movement in time. Then, an additive per-token attribution identifies what the movement is made of, with the vocabulary of two-stage exchange, reimplantation, and debridement receding as single-stage and debridement-with-implant-retention terms rise. An unsupervised geometric decomposition with PCA of mean pooled annual vectors further shows the periprosthetic corpus marching along a data-derived axis running from diagnosis and microbiology toward treatment-procedure content (Kendall τ of year versus the primary axis = +0.94). None of these is individually decisive, and the geometric analysis is exploratory in particular, but their agreement makes the topical shift credible. A potential advantage of a contextual embedding over lexical matching is conceptual robustness to an evolving vocabulary: as a literature matures, the words used for one clinical concept turn over, and a measure built on literal term overlap can decay even when the concept is unchanged, whereas a biomedical embedding may map lexically diverse but conceptually equivalent expressions onto a shared region aligned with the recommendation. This potential advantage was not tested here. In this respect, the method extends the diachronic-embedding tradition by applying time-sliced similarity at the document level against a fixed anchor rather than at the level of individual word senses. [23]

We must also address what TIDE currently fails to properly index. On minimal pairs with identical wording but reversed clinical meaning, neither the embedding nor a lexical baseline changed its score by more than 0.003. The measure therefore tracks topical alignment, not necessarily the evidentiary direction of a claim. The present shortcoming is not only specific to our method but is due to known limitations in the capabilities of text embeddings. Some embedding representations encode a statement and its negation nearly identically, although targeted adaptations can improve this. [24] Identifying whether new evidence supports or opposes a recommendation is a distinct task that TIDE does not yet complete. Being clear about this boundary is important: TIDE can demonstrate that the semantics around a recommendation have been shifting, but that does not necessarily map onto evidentiary trends.

TIDE and its capabilities complement existing automation. Currently, living-evidence and surveillance systems can recall and classify individual study records against human labels. [12,13] However, embedding-derived methods are being applied to guideline work to retrieve literature and rank trials for review updates. [14,15] Unsupervised embeddings have been able to identify how the literature’s themes evolve independently. [16,17] TIDE rather examines, without task-specific supervised training after corpus retrieval, whether the aggregate topical center of gravity of a corpus is moving relative to a recommendation’s own wording over time. Thus, TIDE is best understood as a candidate pre-triage layer that could direct compute- or labor-intensive methods to more probable cases first rather than as a competitor to them. The deterministic, non-generative model design is deliberate because generative models can fabricate and miscite guideline evidence. [21] The auditable and reproducible method presented here avoids entirely importing this genus of failure. [21]

Several limitations bound these conclusions and define the work that remains. We selected the three cases for their documented and clinically expected trajectories, so the results show concordance with known shifts rather than unbiased detection or accuracy. The recovered directions are also sensitive to how each corpus is assembled; we did not establish that they are entirely invariant to the retrieval strategy used to define a corpus, and a topic-only replication in a curated bibliographic source is a necessary next test. The signal is topical rather than evidentiary and is not a predictor of future updates. DRF similarity increased toward all three anchors in the post hoc cross-anchor check, limiting recommendation-specific interpretation. Even an established formal update-signal method reaches only imperfect agreement between predicted update priority and subsequent changes to conclusions(κ = 0.74 across nine reviews). [25] Thus, we frame TIDE as a triage signal whose clinical value remains to be demonstrated. Finally, the analysis rests on a single embedding model, MedCPT, using fixed pretrained encoders across all study years. [20]

The present limitations in TIDE seem surmountable. The natural next step is to move from three curated cases to a systematic evaluation across many recommendations at scale, paired with blinded expert review to test the central untested assumption that topical drift genuinely identifies recommendations that merit reassessment. Complementary directions include replication across multiple encoders and corpora to bound single-model and retrieval sensitivity, and the addition of a deterministic stance-classification layer that could convert a signal that a recommendation’s literature is moving into an estimate of its direction of trend. The authors find the signal demonstrated here credible and interpretable enough to warrant further development and investigation into this direction of recommendation triage.

## CONCLUSIONS

We present TIDE as a deterministic, unsupervised proof of concept examined across three orthopedic recommendations with documented, known trajectories. Using biomedical embeddings, we recovered divergent and convergent signals, with no detectable monotonic trend in ACS that publication and evidence-volume baselines could not capture; in the periprosthetic case, three analyses using different approaches converged on the same interpretable shift. The signal currently tracks topical alignment, not evidentiary direction, a limitation of the present method. TIDE can thus currently flag a recommendation whose literature is moving but cannot itself judge whether the evidence supports or opposes it. The robustness of recovered directions to how each corpus is constructed remains to be fully established. The authors nevertheless find the signal in the known cases credible and sufficiently interpretable to warrant further development and to suggest value in treating embedding representations as an analytical unit rather than solely for retrieval. Our work occupies an intersection of deterministic, anchor-relative, temporal, and unsupervised signal. We intend to generalize and systematize the method for future large-scale analysis.

## Statements and Declarations

### Ethics approval and consent

This study used published bibliographic records, involved no human participants or identifiable patient data, and did not require participant consent.

### Data and code availability

Code, search queries, PMID mappings, frozen vectors and numerical outputs are publicly available at https://github.com/Colbstang/TIDE,revisionbd7586ff8d3cf9f7fd9db769d4e0f42b41f80b07. Article titles and abstracts are excluded from the public repository. Public code reproduces the primary trends and vector-only robustness checks. Text-dependent analyses and exact text-to-embedding reconstruction require the separately retained frozen text archive; access through the corresponding author is subject to applicable rights and permissions.

## Author contributions

**Colby Grames** contributed to study conception, methodology, data acquisition and curation, software development, analysis, validation, visualization, and manuscript drafting and revision.

**Paul Zakarian** contributed to manuscript writing.

**Christopher Franquemont** contributed to research and manuscript review and editing.

**Andrew Cabrera** provided technical input on the methodology.

**Joseph Elsissy** supervised the study and refined the recommendation anchors.

## Ethics statement

The present study used only published and freely available bibliographic data (titles and abstracts) and did not involve human subjects or any potentially identifiable data in any form.

